# Safety and Immunogenicity of CoronaVac and ChAdOx1 Against the SARS-CoV-2 Circulating Variants of Concern (Alpha, Delta, Beta) in Thai Healthcare Workers

**DOI:** 10.1101/2021.10.03.21264451

**Authors:** Nasikarn Angkasekwinai, Jaturong Sewatanon, Suvimol Niyomnaitham, Supaporn Phumiamorn, Kasama Sukapirom, Sompong Sapsutthipas, Rujipas Sirijatuphat, Orasri Wittawatmongkol, Sansnee Senawong, Surakameth Mahasirimongkol, Sakalin Trisiriwanich, Kulkanya Chokephaibulkit

## Abstract

**Importance:** Inactivated vaccine (CoronaVac) and chimpanzee adenovirus-vector vaccine (ChAdOx1) have been more available in resource-limited settings. However, the data comparing between these two vaccines in the same setting are limited.

**Objectives:** To determine adverse events (AEs) and immunogenicity of CoronaVac and ChAdOx1 in health care workers (HCWs).

**Design:** This prospective study was conducted from February to July 2021.

**Setting:** A single center, university-based tertiary care center in Bangkok.

**Participants:** Healthy HCWs.

**Exposure:** Two doses of CoronaVac (4 weeks apart) or ChAdOx1 (8 weeks apart) intramuscularly.

**Main Outcomes and Measures:** Self-reported AEs were collected for 7 days following each vaccination using electronic diary. The immunogenicity was determined by the level of IgG antibodies against receptor binding domain (RBD) of the SARS-CoV-2 spike protein (S1 subunit). The 50% plaque reduction neutralization tests against original Wuhan strain and circulating VOCs were performed in subset of samples at 2 weeks after the second dose.

**Results:** Of the 360 HCWs, 180 received each vaccine. The median (interquartile range: IQR) age was 35 (29-44) years old and 84.2% were female. Participants who received ChAdOx1 reported higher frequency of AEs than those received CoronaVac after both the first dose (84.4% vs. 66.1%, *P* < 0.001) and second dose (75.6% vs. 60.6%, *P* = 0.002), with more AEs in those younger than 30 years of age for both vaccines. The seroconversion rate was 75.6% and 100% following the first dose of CoronaVac and ChAdOx1, respectively. All participants seroconverted at 2 weeks after the second dose. The anti-SARS-CoV-2 RBD IgG levels induced by CoronaVac was lower than ChAdOX1 with geometric means of 164.4 and 278.5 BAU/mL, respectively (*P* = 0.0066). Both vaccines induced similar levels of neutralizing antibodies against the Wuhan strain, geometric mean titer (GMT) of 337.4 vs 331.2; however, CoronaVac induced significantly lower GMT against Alpha (23.1 vs. 92.5), Delta (21.2 vs. 69.7), and Beta (10.2 vs. 43.6) variants, respectively.

**Conclusions and Relevance:** CoronaVac induces lower measurable antibodies but with lower frequency of AEs than ChAdOx1. The low neutralizing antibodies against the circulating VOCs induced by CoronaVac supports the need for earlier boosting to prevent breakthrough infections.

**Trial Registration:** TCTR20210720002 https://www.thaiclinicaltrials.org/

**Question:** What is the difference between CoronaVac and ChAdOx1 vaccines on safety and immunogenicity against the circulating variants of concern (VOCs) in the same setting?

**Findings:** This prospective study in 360 healthy health care workers reported higher frequency of adverse events following ChAdOx1 than CoronaVac particularly in those younger than 30 years old. The ChAdOx1 induced 3.3-4.3 times higher neutralising antibodies against VOCs than CoronaVac.

**Meaning:** The 2-dose CoronaVac vaccination induced significantly lower level of neutralizing antibody against the circulating VOCs. An earlier booster may be needed to prevent breakthrough infection.

## Introduction

Vaccination has been the most effective measure to control the outbreaks of coronavirus diseases-2019 (COVID-19). Although the immunogenicity and efficacy reported in phase III of different vaccines varied, all vaccines that passed the WHO Emergency Use Listing Procedure (EUL) have been shown to prevent severity and mortality of COVID-19. Due to its urgency, the data available at the stage of approval for emergency use are generally limited. The adverse events in various settings as well as rare adverse events are unknown.^1-4^

Most resource-limited settings had limited access to vaccines in early 2021. In most part of Asia, including Thailand, the whole-cell inactivated vaccines such as CoronaVac (*Sinovac, Life Sciences*) and chimpanzee adenovirus-vector (ChAdOx1) vaccine (AZD1222, *AstraZeneca*/*Oxford*) were available for procurement earlier than other vaccines. The phase III trials indicated 76% efficacy of ChAdOx1 and 50.8% efficacy of CoronaVac in preventing symptomatic infection, and both vaccines effectively prevented severe diseases with 100% efficacy.^5-7^ Since the studies were conducted in different settings and with limitations, the efficacy in real world may differ from the trials due to heterogeneity of the population receiving the vaccines. The variants of interest (VOIs) and concern (VOCs) that are impacting on transmissibility and disease severity emerged during the late 2020, posing the risk of breakthrough infections, either after natural infections or after vaccination.^8, 9^ A previous study found three to five fold lower neutralizing titers against the Alpha (B1.1.7) and Delta (B1.1617.2) variants compared with the original strain.^10^ In many settings, the Alpha variant was predominant in April 2021, but has since been taken over by the Delta variant in August 2021. The Beta variant has also emerged in many areas, but, like the other VOCs, it has been generally out-competed by the more infectious Delta. The appearance of these VOCs raised the concern of reduced effectiveness of the COVID-19 vaccines that are widely used.

Healthcare workers (HCWs) are the priority population for COVID-19 vaccination due to an increased risk of exposure to SARS-CoV-2.^11^ To ensure the efficacy and safety of the COVID-19 vaccines, CoronaVac and ChAdOx1, which have been widely used in many settings, we investigate the adverse events and immunogenicity following the vaccination of these two vaccines in the same setting including the comparison with convalescent sera from natural infected individuals.

## Material and Methods

### Study design and participants

This single-center prospective cohort study enrolled 360 healthy HCWs aged 18 years or older at Siriraj Hospital, a university-based referral center located in Bangkok, Thailand from February to July 2021. The participants were excluded from the study if they had the following conditions: confirmed SARS-CoV-2 infection, had received current prophylactic treatment or investigational agents against COVID-19 within 90 days, had unstable underlying diseases that may compromise the immune responses, had a history of vaccine hypersensitivity, were pregnant, were immunocompromised or receiving immunosuppressive agent. The study protocol was approved by the Siriraj Institutional Review Board (COA no. Si 171/2021).

### Study Procedures

After providing written informed consent, the participants were randomly assigned to receive either two doses of CoronaVac at 4 weeks interval or two doses of ChAdOx1 at 10 weeks interval. The HCWs aged 60 years or older were assigned to ChAdOx1 based on the local guidelines at that time. The participants were observed for at least 30 minutes following the vaccination for any immediate adverse events. The participants were instructed to submit self-assessment report using an electronic diary (eDiary) in the Google Form for seven days after each dose of vaccination for any adverse events (AEs) both solicited local and systemic reactions. The solicited local AEs include pain, erythema, and swelling/induration at the injection site, and localized axillary swelling or tenderness ipsilateral to the injection arm. The systemic AEs include headache, fatigue, myalgia, arthralgia, diarrhea, dizziness, nausea/vomiting, rash, fever, and chills. The severity of solicited AEs was graded using a numerical scale from 1 to 4 based on the Common Terminology Criteria for Adverse Events - Version 5.0 guide by the United States National Cancer Institute (NCI/NIH).

#### Chemiluminescent microparticle assay (CMIA) for anti-SARS-CoV-2 RBD IgG and anti-NP

Blood samples were collected at baseline (pre-dose 1), on the day of second dose (pre-dose 2), 4 weeks after the second dose and 8-12 weeks after the second dose. A subgroup of participants who received ChAdOx1 were randomly invited to test for anti-SARS-CoV-2 RBD IgG at 4 weeks after the first dose. The plasma samples were isolated from the blood collected in tubes with sodium citrate solution and stored at -80ºC. The level of antibody response (IgG) against receptor binding domain (RBD) of the SARS-CoV-2 spike protein (S1 subunit) was determined by a CMIA using the SARS-CoV-2 IgG II Quant (Abbott, List No. 06S60) on the ARCHITECT i System The anonymous convalescent sera of mildly symptomatic infection from outbreaks in Thailand caused by B.1.36.16 (D614G) strain in late December 2020 collected at 4 or 12 weeks of illness were tested as the reference. This assay linearly measures the level of antibody between 21.0 - 40,000.0 arbitrary unit (AU)/mL, which was converted later to WHO International Standard concentration as binding antibody unit per mL (BAU/mL) following the equation provided by the manufacturer (BAU/mL = 0.142 x AU/mL). The level greater or equal to the cutoff value of 50 AU/mL or 7.1 BAU/mL was defined as seropositive. Seroconversion was determined as becoming seropositive in those who had seronegative at baseline or an increase of 4-fold titers above the baseline seropositive levels.

Qualitative antibody response against the SARS-CoV-2 nucleocapsid protein (NP) in the plasma samples was determined by CMIA using the SARS-CoV-2 IgG (Abbott, List No. 06R86) on the ARCHITECT i System at baseline and 4 weeks after the second dose.

#### 50% Plaque reduction neutralization test (PRNT_50_) against SARS-CoV-2 strains

Subgroup of subjects in each vaccine group were randomly invited for additional blood collection at two weeks after the second dose for determining the level of anti-SARS-CoV-2 RBG IgG and the neutralizing antibody titers against original (Wuhan) strain and VOCs, which were Alpha (B1.1.7), Delta (B1.1617.2), and Beta (B.1.351) strains by 50% plaque reduction neutralization test (PRNT_50_). The convalescent sera of the mildly symptomatic patients recovered from D614G strain infections in late December 2020 and Delta strain in June 2021, collected by the Ministry of Public Health (MOPH) at 2 weeks of illness, were tested for PRNT50 against Delta variant as reference. Any titers below the detection limit of PRNT_50_, which is 1:10 will be indicated as an average of 1:5 for statistical analyses. Briefly, Vero cells were seeded at 2×10^5^ cells/well/3 mL and placed in 37°C, 5% CO_2_ incubator for 1 day. Sera were serially diluted at 1:10, 1:40, 1:160 and 1:640, respectively. The SARS-CoV-2 virus was diluted in culture medium to yield 40-120 plaques/well and mixed in equal volume of the diluted serum at 37°C in water bath for 1 hour. Convalescent and uninfected sera were used as assay controls. The virus-serum mixture (200 μL) was inoculated into Vero-cell monolayer in triplicate and the culture plates were rocked every 15 minutes for 1 hour. Three mL of the overlay semisolid medium containing 1% of carboxymethylcellulose (Sigma Aldrich, USA), 1% of 10,000 units/mL Penicillin with 10,000 ug/mL Streptomycin (Sigma, USA) and 10% fetal bovine serum (FBS) were added to the cells after removing the virus-serum mixture. Cells were incubated at 37°C, 5% CO_2_ for 7 days, then fixed with 10% (v/v) formaldehyde and stained with 0.5% crystal violet in phosphate-buffered saline (PBS). The number of plaques was counted and the percentage of PRNT_50_ was calculated. The titer of each sample was defined as the reciprocal of the highest test serum dilution, which the virus infectivity was reduced by 50% of an average plaque counts in the virus control wells and was calculated by using a four-point linear regression method.

### Statistical Analysis

The AEs endpoints were presented as frequencies and the Chi-square test was used to test for statistical difference. The immunological endpoints including the level of anti-SARS-CoV-2 RBD IgG and PRNT_50_ titer were reported as geometric mean (GM) with 95% confidence interval (CI). The PRNT_50_ titer below 10 were arbitrarily assigned a value of 5. GraphPad Prism 9 version 9.2.0 (283) (GraphPad Software, CA, USA) was used for unpaired t-test analyses to compare GM of the IgG concentrations between groups and Pearson‘s correlation coefficient to assess the correlation between log10 of anti-SARS-CoV-2 RBD IgG and log10 of PRNT_50_. The ANOVA was performed to examine the geometric mean of anti-SARS-CoV-2 RBD IgG among different age groups using STATA version 17 (StataCorp, LP, College Station, TX, USA).

## Results

Of the 360 HCWs enrolled (Figure 1), 180 were in each vaccine group with the median (IQR) age of 35 (29-44) years old, and 303 (84.2%) were female. There were 14 participants (3.9%) aged 60 years or older and all received ChAdOx1 vaccine. The participants who received ChAdOx1 were older than those who received CoronaVac with the median of 40 (32-48) years old and 31 (26-39) years old, respectively (*P* < 0.001). Participants who received ChAdOx1 had higher frequencies of hypertension (10% vs. 2.2%, *P* = 0.003) and dyslipidemia (10.6% vs. 2.8%, *P* = 0.005) than those received CoronaVac (Table 1).

**Figure 1.**
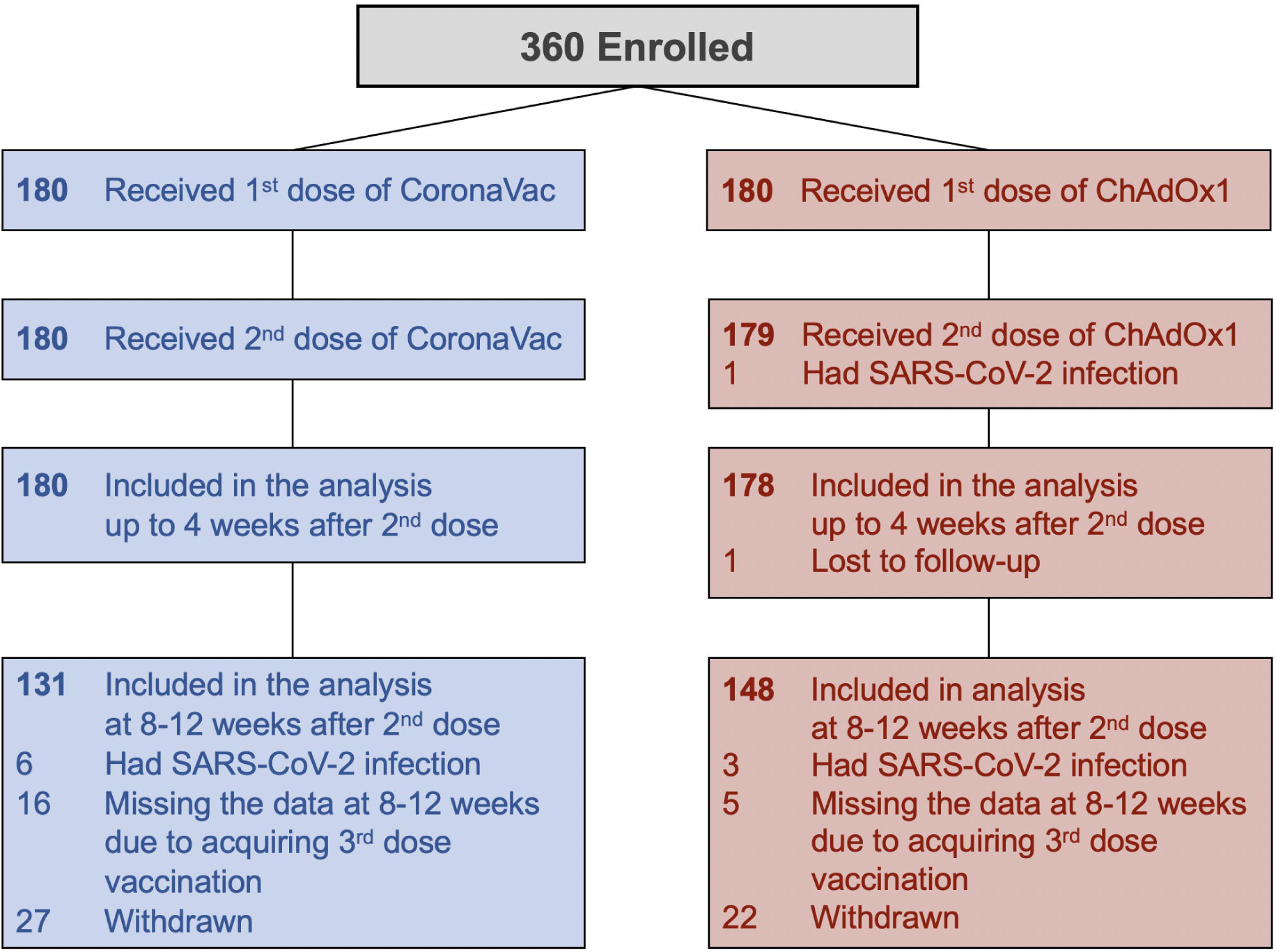
Flow diagram of enrollment and vaccination of healthcare worker participants.

**Table 1:** Baseline characteristics of 360 health care worker participants by vaccine type.

|  | Total | Vaccine Group |  | *p*-value |
| --- | --- | --- | --- | --- |
|  | n=360 | ChAdOx1 (n=180) | CoronaVac (n=180) |  |
| Male, n (%) | 57 (15.8) | 30 (16.6) | 27 (15.0) | 1.000 |
| Age: year (median, IQR) | 35.00 (29.00, 44.00) | 40.00 (32.00, 48.00) | 31.00 (26.00, 39.00) | <0.001* |
| Weight: kilogram (median, IQR) | 59.80 (52.43, 69.38) | 59.05 (53.13, 69.30) | 60.00 (52.10, 69.55) | 0.945 |
| Body mass index: kg/m<sup>2</sup> (median, IQR) | 23.12 (20.50, 26.36) | 23.26 (20.70, 26.29) | 22.87 (20.37, 26.36) | 0.656 |
| Comorbidity |  |  |  |  |
| Hypertension, n (%) | 22 (6.1) | 18 (10.0) | 4 (2.2) | 0.003* |
| Dyslipidemia, n (%) | 24 (6.7) | 19 (10.6) | 5 (2.8) | 0.005* |
| Diabetes mellitus, n (%) | 9 (2.5) | 6 (3.4) | 3 (1.7) | 0.337 |
| Obesity, n (%) | 7 (1.9) | 4 (2.2) | 3 (1.7) | 1.000 |

\*Statistical significance (*p*-value < 0.05) comparing between ChAdOx1 and CoronaVac group.

### Adverse Events

Subjects who received ChAdOx1 reported any AEs more frequent than CoronaVac recipients following the first and second dose vaccinations (84.4% vs. 66.1%, *P* < 0.001 and 75.6% vs. 60.6%, *P* = 0.002, respectively), (Figure 2A, B). All AEs were mild (grade 1) or moderate (grade 2) in severity and recovered within 2-3 days. Compared with the first dose, the ChAdOx1 vaccine group reported less frequent and milder AEs following the second dose, while the CoronaVac vaccine group reported overall similar frequency of systemic AEs between the first and second dose, but with more local AEs after the second dose. There was no serious AEs reported in any participant. In both vaccine groups, the subjects younger than 30 years of age reported more systemic AEs, particularly myalgia and fever (Figure 2 C, D). There was no difference between gender for overall AEs after the CoronaVac and after the first dose of ChAdOx1; however, female reported overall AEs more than male (70.3% vs. 56.1%, *P* = 0.035) after the second dose of ChAdOx1.

**Figure 2.**
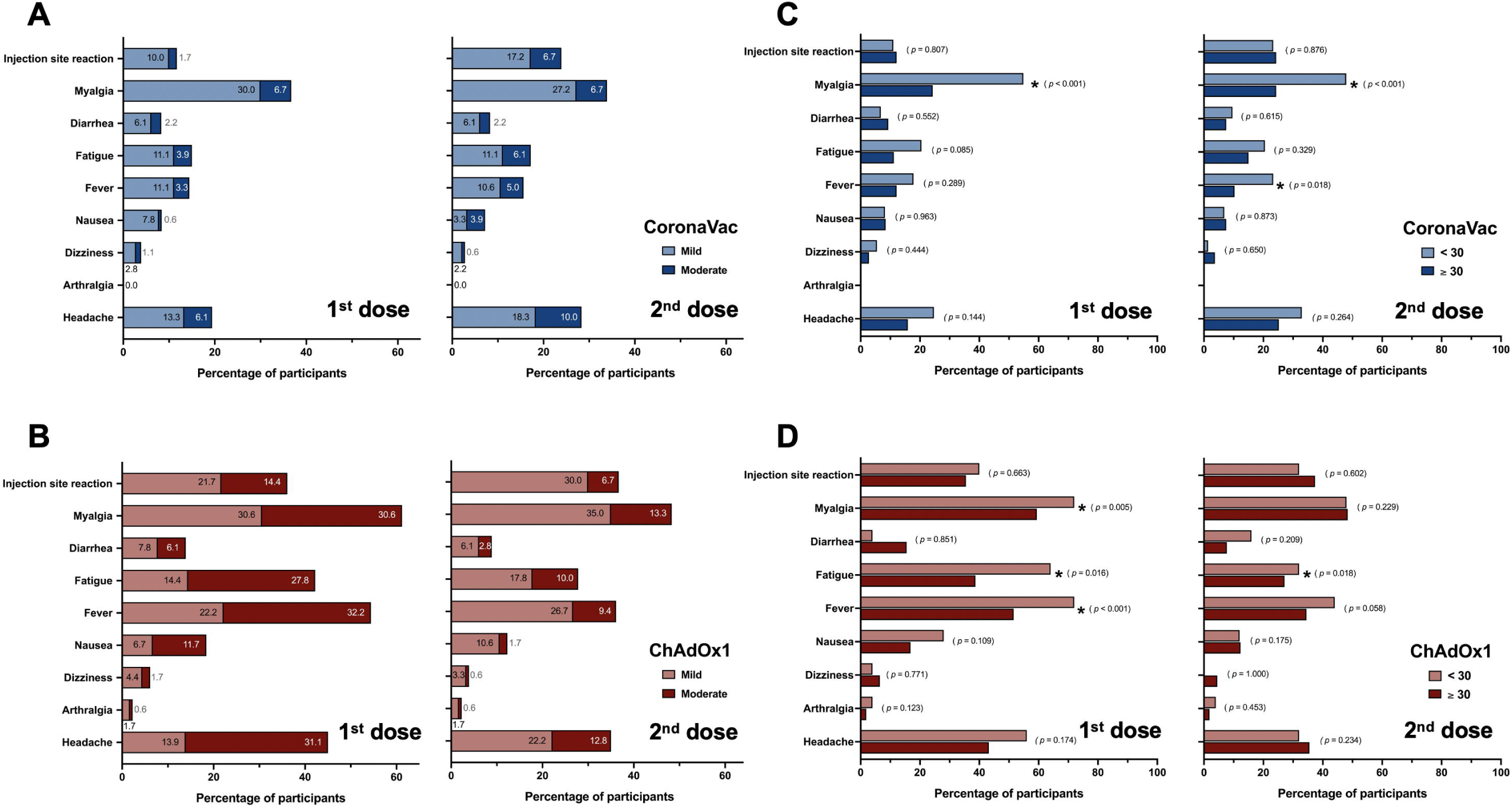
Adverse events following CoronaVac or ChAdOx1 vaccination. The stacked bars showed the percentage of participants who had indicated mild and moderate adverse events after the first and second dose of CoronaVac (A) and ChAdOx1 (B). The stacked bars showed the percentage of participants aged lower or above thirty years old, who had indicated adverse events after the first and second dose of CoronaVac (C) and ChAdOx1 (D). Chi-square was used for statistical analyses and the *p* values were shown on the graphs.

### Immunogenicity

All participants were negative for anti-SARS-CoV-2 NP IgG at baseline and none had history of COVID-19. At 4 weeks after the first dose, CoronaVac induced a lower anti-SARS-CoV-2 RBD IgG seroconversion rate than ChAdOx1 (75.6% (136/180) vs 100% (180/180), with lower antibody levels (geometric mean (GM) 12.7 BAU/mL vs. 69.8 BAU/mL). For the ChAdOx1 recipients, the level of anti-SARS-CoV-2 RBD IgG decreased to a GM of 37.1 BAU/mL prior to the second dose vaccination. After the second dose, both vaccines induced 100% seroconversion within 2 weeks. ChAdOx1 induced higher levels of anti-SARS-CoV-2 RBD IgG than CoronaVac at all follow-up time points after the second dose: 2 weeks, GM 278.5 BAU/mL vs. 164.4 BAU/mL (*P*= 0.0066); 4 weeks, GM 178.2 BAU/mL vs. 94.8 BAU/mL (*P* < 0.0001); and 8-12 weeks, GM 92.5 BAU/mL vs. 34.7 BAU/mL (*P* < 0.0001). In comparison with the D614G convalescent sera at 4 and 12 weeks, CoronaVac induced significantly lower levels of anti-SARS-CoV-2 RBD IgG while ChAdOx1 induced higher levels (Figure 3A).Among those who received ChAdOx1 vaccine, the anti-SARS-CoV-2 RBD IgG levels at 4 weeks after the second dose varied by age group (*P* < 0.0001 by ANOVA) with a trend towards higher levels in younger age groups. This age-related response was not seen in CoronaVac group (*P* = 0.058, Figure 3B).

**Figure 3.**
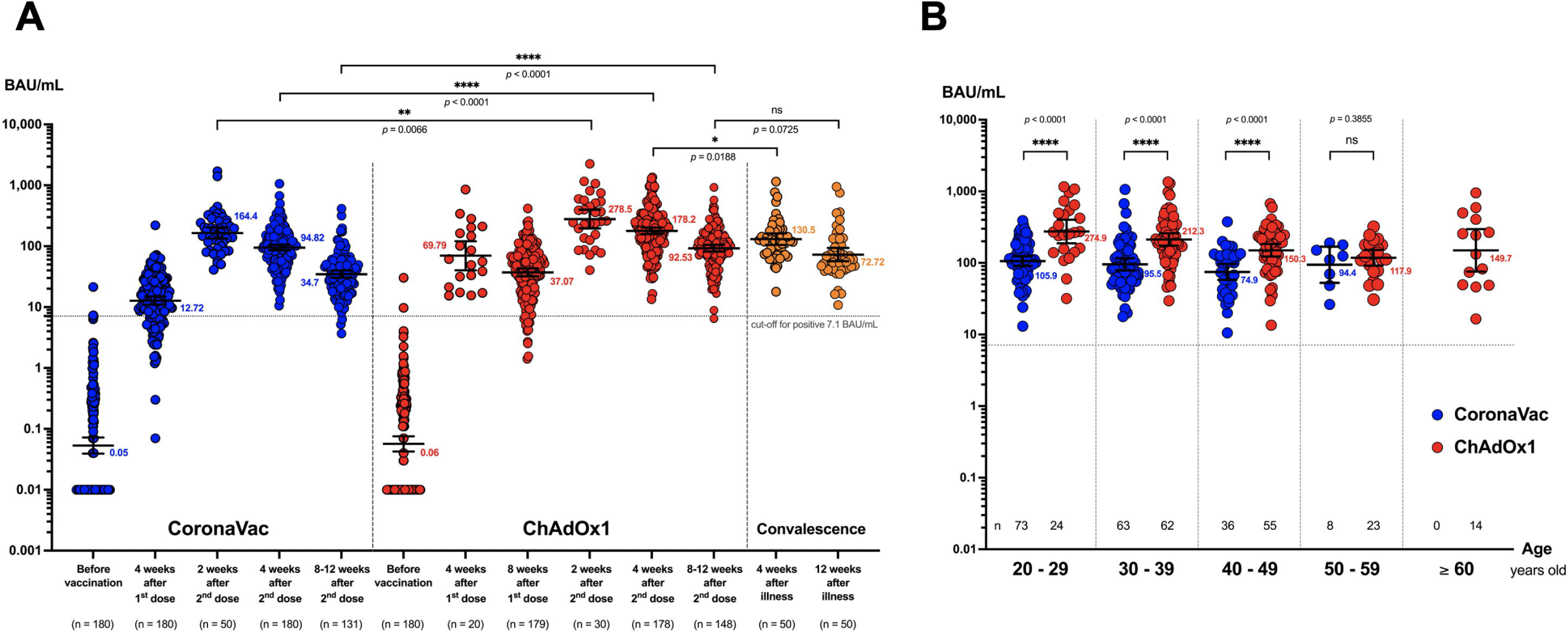
Anti-SARS-CoV-2 RBD IgG levels following CoronaVac or ChAdOx1 vaccination. (A) Anti-SARS-CoV-2 RBD IgG levels in the plasma of study subjects before and various time points after vaccination with CoronaVac (blue) or ChAdOx1 (red). Convalescent sera at 4 weeks and 12 weeks after illness from the patients who recovered from the COVID-19 during Dec 2020 and early 2021 were included as reference level (orange). (B) Anti-SARS-CoV-2 RBD IgG levels in the plasma of study subjects among different age groups at 4 weeks after the second dose vaccination of CoronaVac (blue) or ChAdOx1 (red). The numbers in the graph represent geometric mean and the error bars represent 95% confidence interval. The number of tested samples were indicated below each time point. Two-tailed unpair *t* test was used to compared the IgG level between two conditions with indicated *p* value.

In addition, CoronaVac induced anti-SARS-CoV-2 NP IgG seroconversion in 53% of vaccinees at 4 weeks following the second dose. Individuals seropositive for SARS-CoV-2 NP IgG had significantly higher level of anti-SARS-CoV-2 RBD IgG compared to those seronegative for SARS-CoV-2 NP IgG with the GM of 121.7 BAU/mL and 71.3 BAU/mL, respectively (*P* < 0.001), (Supplementary Figure S1). None but one subject who received ChAdOx1 had anti-SARS-CoV-2 NP IgG seroconversion; the exceptional subject had mild symptomatic SARS-CoV-2 infection approximately 4 weeks after the first dose.

The GM titers (GMT) of the PRNT_50_ against the original Wuhan strain and VOCs at 2 weeks following the second dose vaccination was shown in Figure 4A. Both CoronaVac and ChAdOx1 induced similar GMT against the original Wuhan strain (337.4 vs 331.2, *P* = 0.736). Compared with CoronaVac, the ChAdOx1 induced significantly higher GMTs against all VOCs, (Alpha 92.5 vs. 23.1, Delta 69.7 vs. 21.2, and Beta 43.6 vs. 10.2, *P* < 0.001 for all comparisons), (Figure 4A). The PRNT_50_ GMTs against Delta variant induced by CoronaVac were significantly lower than convalescent sera of individuals infected with D614G strain (21.2 vs 96.6, *P* <0.0001), while ChAdOx1 induced similar level (*P* = 0.183). The convalescent sera of individuals infected with the Delta variant induced GMT of 490.1, which was significantly higher than the GMTs induced by both vaccine groups (Figure 4B).

**Figure 4.**
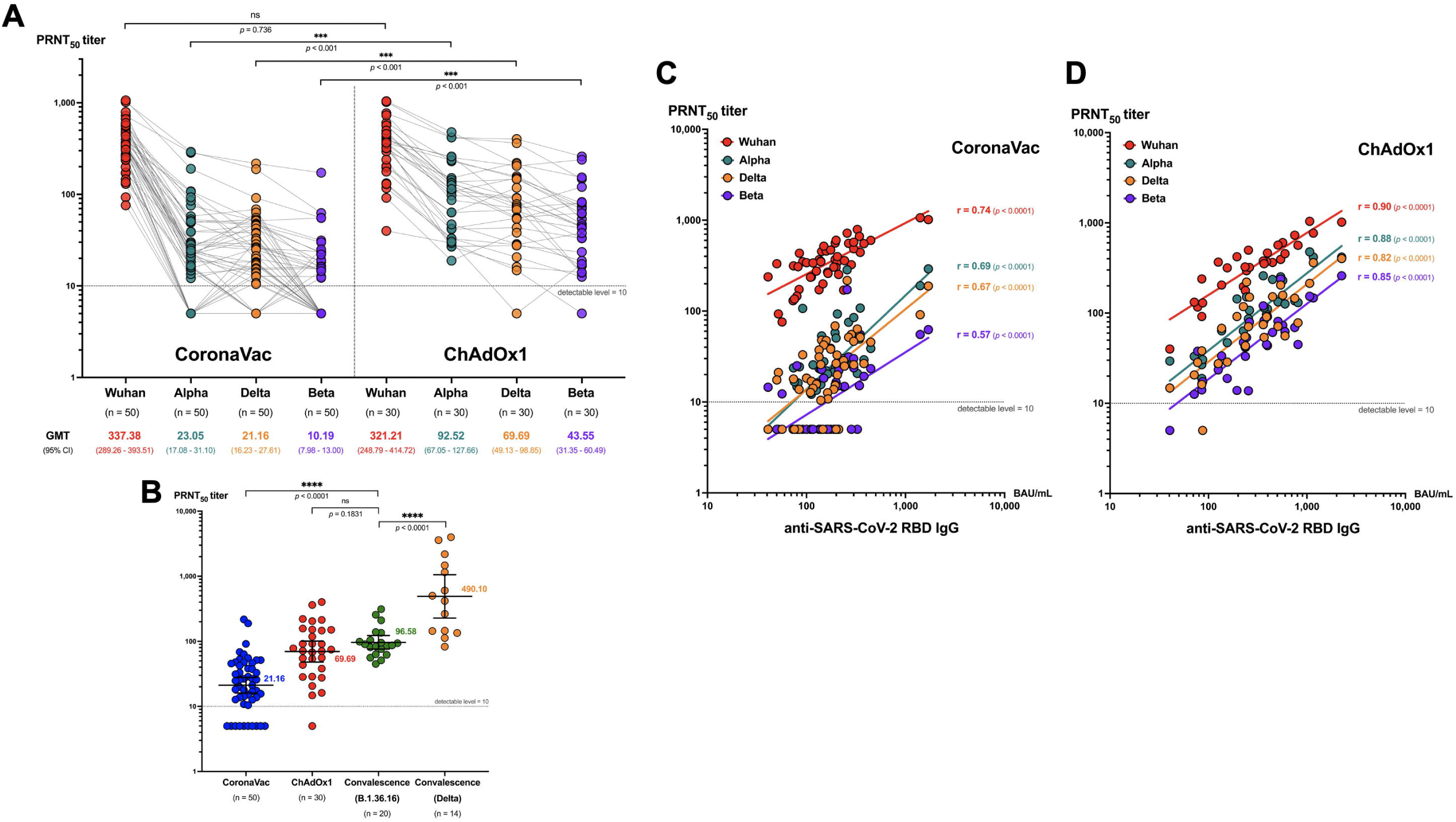
Plaque reduction neutralization test (PRNT_50_) titers for different SARS-CoV-2 strains. (A) Dot plots demonstrates PRNT_50_ titer against Wuhan (red), Alpha (teal), Delta (orange) and Beta (purple) strains in the plasma of study subjects at 2 weeks after two doses of CoronaVac or ChAdOx1. (B) Scatter dot plots demonstrates PRNT_50_ titer against delta strain at 2 weeks after 2-dose vaccination with CoronaVac (blue) and ChAdOx1 (red) compared with the convalescent sera at 2 weeks after illness of the patients infected with Delta strain (orange) and B.1.36.16 strain (green). The PRNT_50_ titer of 1:5 was used for all that were below the detectable level (< 1:10). The geometric mean titer (GMT) and lower and upper 95% confidence interval (CI) are indicated. (C) Correlation between the level of anti-SARS-CoV-2 RBD IgG and plaque reduction neutralization test (PRNT_50_) titers for wildtype and VOC. Dot plots shows the correlation between the level of anti-SARS-CoV-2 RBD IgG and PRNT50 titer against Wuhan (red), alpha (teal), delta (orange) and beta (purple) strains in the plasma of study subjects at 2 weeks after two doses of CoronaVac (n = 50) or (D) ChAdOx1 (n=30). The PRNT_50_ titer of 1:5 was used for all that were below the detectable level (< 1:10). Pearson‘s correlation coefficient (r) with *p* value for each strain was indicated.

The anti-SARS-CoV-2 RBD IgG levels and the PRNT_50_ titers against all four strains were strongly correlated in the ChAdOx1 group (r = 0.82 to 0.9, *P* < 0.0001). In contrast there was only moderate correlation in the CoronaVac group (r = 0.57-0.74, *P* < 0.0001) (Figure 4C, D).

## Discussion

Our study provided important knowledge about AEs, immunogenicity and dynamics of the immune responses induced by CoronaVac and ChAdOx1 in the same population at the same time. We also evaluated the neutralizing antibody titers induced by the vaccines against circulating VOCs. This study is relevant to many settings where these two vaccines are widely used, especially during outbreaks caused by VOCs.

We found both vaccines to be well tolerated with more AEs following ChAdOx1 than CoronaVac. Any AEs following the first dose of ChAdOx1 were reported at a lower rate (66.1%) in our study than those reported in the phase III trial (86% in participants age 18-55 years)^12^, but closer to the rate of 59% reported in the United Kingdom (UK) where a self-reporting application was used.^13^ Systemic AEs for both vaccines were significantly higher in the vaccinees younger than 30 years of age with more AEs reported by females following the second dose. This was consistent with the self-report application in the UK. For CoronaVac, the AEs rates were similar to those reported in a phase II trial in China and a phase III trial in Brazil^6, 7^, but the higher AE rate in younger individuals has not been reported.

Overall, the anti-SARS-CoV-2 RBG IgG induced by the ChAdOx1 was significantly higher than that the levels induced by the CoronaVac at all time points up to 8-12 weeks following the second dose of vaccination. The PRNT_50_ GMT induced by the CoronaVac and the ChAdOx1 were similar against the Wuhan strain, but the ChAdOx1 induced 3-4 times higher GMTs against each VOCs than CoronaVac. Compared with those of the Wuhan strain, the PRNT_50_ GMTs against the VOCs were 16-30 times lower in the CoronaVac group and 5-7 times lower in the ChAdOx1 group. The PRNT_50_ GMTs against the Alpha and Delta variants were not different in both vaccine groups and were much higher than those against the Beta variants. These data concurred with the previous study and suggested more distance genetic variability of the Beta variant.^14^ This suggest that both vaccines are less effective against the Beta variant.

The neutralizing antibodies induced by both vaccines against the Delta variant were lower than convalescence sera from post Delta variant infection, with lower levels in the CoronaVac group as seen in a previous study. ^15^ Although protective levels of the neutralizing antibodies have not yet been established, Khoury et al. suggested the protective threshold to be at 28.6% (95% CI□ = 19.2–29.2%) of the mean convalescent level.^16^ Using this assumption, based on our data, both vaccines would not provide effective protection against the Delta variant, but breakthrough VOC infections would be more likely in those who had received CoronaVac.

The presence of anti-SARS-CoV-2 NP antibody is used as the marker for natural infection and is expected to be positive following whole virus inactivated vaccine. However, we found positive anti-SARS CoV-2 NP antibody in only about half of the CoronaVac recipients, consistent with a previous report^17^. This could suggest that CoronaVac is not as immunogenic as natural infection.

After completing 2 doses of both vaccines, the level of anti-SARS CoV-2 RBD IgG in the blood peaks at 2 weeks before declining by 1.5 to 1.7 times at 4 weeks and 3 to 5 times at 8-12 weeks afterward. This half-life is shorter than that suggested for the half-life of neutralizing antibody of 108 days.^16^ Our data also suggests that the peak anti-SARS-CoV-2 RBD IgG antibody response is best measured around 2 weeks after completing the 2-dose schedule. Consistent with the previous study^18^, we found a strong positive correlation of neutralizing antibodies and anti-SARS-CoV-2 RBD IgG, supporting the use of anti-SARS-CoV-2 RBD IgG as a proxy marker for neutralizing antibodies.

This study has several limitations. Although the strength of this study is the homogeneity of the population, the population included only healthy HCWs, which may not be able to generalize to other population. In addition, only a subset of participants with small sample size were tested for the PRNT_50_ against the VOCs; however, the difference was able to be demonstrated. This study also lacks the data of cell-mediated immune responses, which could be another important factor to control the severity of viral infection. This study adds to the limited data available for CoronaVac and ChAdOx1 in the Asian population and provides direct comparison of these two vaccines particularly against the VOCs by using the gold-standard measurement of neutralizing antibody.

## Conclusion

CoronaVac and ChAdOx1 are well tolerated vaccines and equally immunogenic for original Wuhan strain. However, the ChAdOx1 induced more AEs (mild to moderate), but provide higher level of anti-SARS-CoV-2 RBD IgG and the PRNT_50_ against all VOCs compared with the CoronaVac. Both vaccines induced neutralizing antibodies against the Delta variant but at a significantly lower level than titers detected in the Delta convalescent sera and probably insufficient to prevent infection. These results support the need for booster vaccination to prevent breakthrough infections from the VOCs, particularly among those who received the CoronaVac.

## Data Availability

Not Available

## Acknowledgement

The authors gratefully acknowledge the Siriraj Institute of Clinical Research (SICRES) team, Abbott Laboratories Ltd. for technical supports and all health care workers who took part and enabled this study to be possible. We thank Professor Kim Mulholland and Zheng Quan Toh at Murdoch Children‘s Research Institute for their critical data review and suggestion.

## Figure legends

**Supplementary Figure S1.**
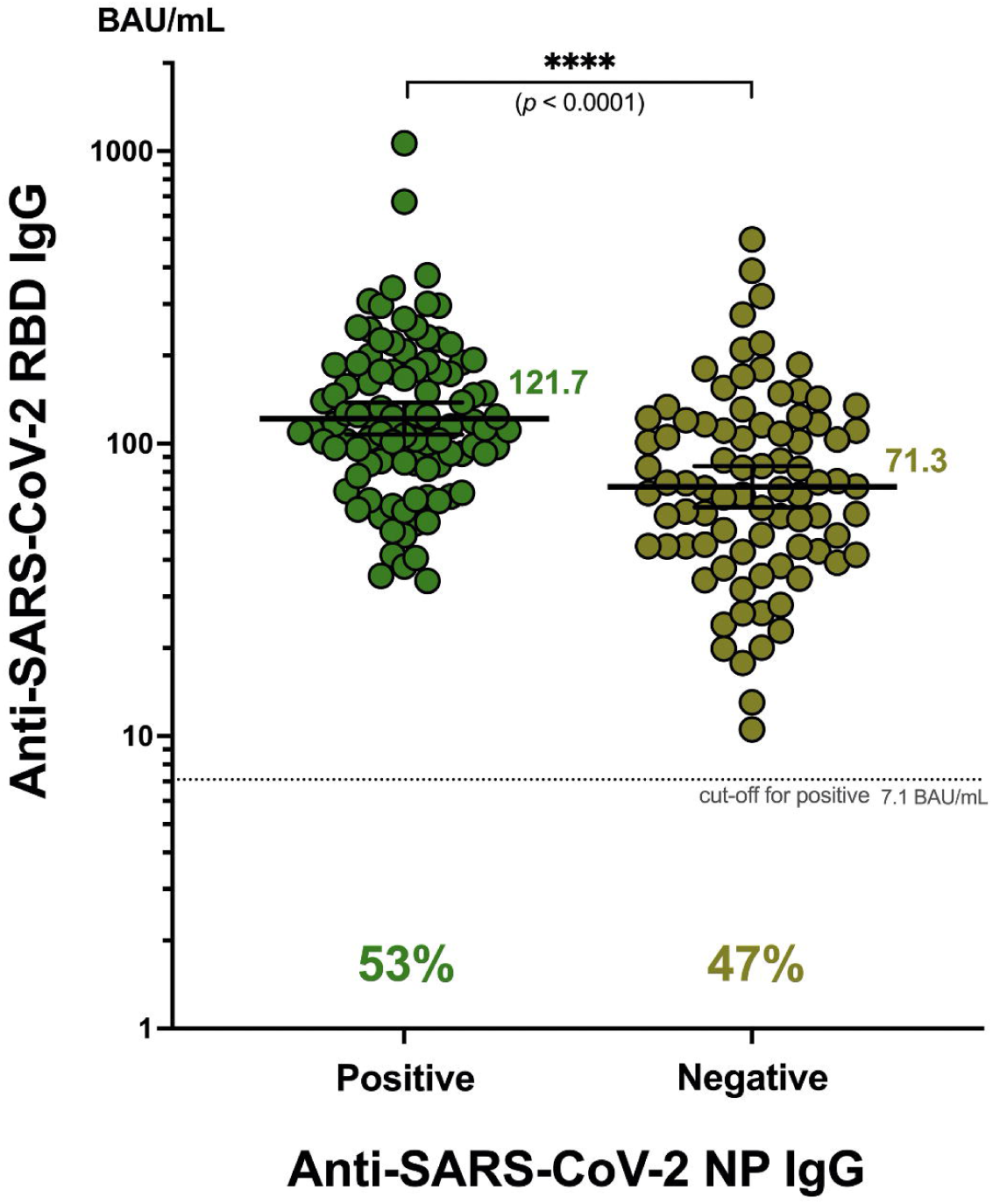
Correlation of anti-SARS-CoV-2 RBD IgG and anti-SARS-CoV-2 nucleocapsid protein antibody in the subjects who received CoronaVac (n=180).

